# Study on the value of isolation and extraction of pathological positive cells to improve the detection rate:control trial

**DOI:** 10.1101/2024.12.11.24318866

**Authors:** JianQuan Luo, JiaCheng Yang

## Abstract

**OBJECTIVE:** The density of pathological positive cells was calculated by flow cytometry, and the target cells were isolated and extracted to improve the detection rate.

**DESIGN:** a case-control study.

**SETTING:** Hematology Department of Hospital

**PARTICIPANTS:** Selected from MDS 33 cases, 17 cases of malignant tumors, 29 cases of lupus erythematosus, 4 cases of plasmodium falciparum disease, a total of 76 cases as positive blood specimens.

**MAIN OUTCOME MEASURES:** Design Research group (group I) : Flow cytometry was used to analyze and calculate the density of various cell types, and then cell separation solution was used to accurately separate and extract target cells of the same density by adjusting the gradient of two different densities, that is, to collect pathologically positive blood cells of the same density. A control group (group II) was established: the whole blood samples of the same patient were compared and analyzed by direct push-film observation.

**RESULTS:** As shown in Figure 2AB-3AB-4AB-5AB, A large number of positive abnormal cells were extracted from group I, which improved the accuracy and specificity of diagnosis. In the control group (group II), only a few positive abnormal cells were found, which was very easy to misdiagnose and miss diagnosis. Data are analyzed from Table 3. According to the statistical results of group I, the positive detection rates were MDS=76%, malignant tumor cells =53%, lupus erythematosus cells =62%, Plasmodium falciparum =75%. In the control group (group II), the positive detection rate was MDS=27%, malignant tumor cells =11%, lupus erythematosus cells =33%, Plasmodium falciparum =25%. Chi-square test was used to compare the mean value of independent samples between groups, and the difference was statistically significant (P<0.05).

**CONCLUSIONS:** The density of pathologically positive blood cells can be measured by flow cytometry according to the specimens that need to be reviewed by microscopy as indicated by blood routine, and the target cells extracted can be accurately separated, that is, the same density of pathologically positive blood cells can be collected. Combined with the gold standard method of artificial microscopy, the detection rate was significantly improved, and the occurrence of misdiagnosis and missed diagnosis was reduced. For the difference in cell density of various forms in different types of specimens, it only needs to be distinguished from normal cell density, and abnormal pathological positive cells can be accurately extracted, which can help diagnose the positive detection rate of diseases, which is of great significance and value

## Introduction

When a person develops symptoms of ill health, the first doctor will first consider doing routine blood tests to help analyze and diagnose the patient’s health condition. In order to improve the diagnostic value of blood routine reports, in 2005, the International Consensus Expert Hematology Evaluation recommended 41 rules for the analysis of blood cells [1], aiming to prevent misdiagnosis and missed diagnosis of instrument measurement results.

Flow cytometry can detect various physical parameters of cells [2] and can analyze and calculate specific cell population density.

According to the blood routine automatically prompts the need to review the blood film observation review of blood samples,The density of various types of cells was measured and calculated by flow cytometry, and then the target cells with the same density were accurately separated and extracted by adjusting the gradient of two different densities with cell separation solution, that is, the same density of pathologically positive blood cells were collected. The detection rate of pathology positive cells in blood smear was improved rapidly.

## Methods

In this study,meet the same standards for ethics of experimentation and research integrity as the research study. proof of ethics approval:Approval Number (ID Number): (Medical Review 2023 No. 61) 2023-061

This technique is used to extract target cells from blood samples of the same patient, analyze and diagnose the positive detection rate of pathological cells, and compare the detection rate with that of whole blood samples directly and automatically, so as to evaluate the application value of the technology of measuring and extracting pathological positive cells. The results of this study are reported as follows:

## 1. Materials and Methods

### 1.0 Materials and reagents

1.1 From January 2021 to December 2023 in our hospital, 33 patients with MDS, 17 patients with malignant tumor, 29 patients with SLE, and 4 patients with plasmodium infection were selected. A total of 76 patients with positive blood samples were included in the study group (group I). The diagnosis of confirmed cases is based on the Chinese Hematology (6th Edition) standard [3]. In the control group (group II), the whole blood samples of the same patients were applied to the automatic blood routine assembly line in the laboratory department of the hospital (result data analysis of Mindray CAL8000 automatic slide reading system) [4]. Abbreviated note [5] : myelodysplastic syndromes (MDS), malignant tumors are cases that can metastasize tumor cells in the blood, Systemic Lupus Erythematosus (SLE), Plasmodium-infected patients are malaria patients infected inside the red blood cells.

### 1.2 Use of reagents, principles, steps

1.2.1 Follow the complete steps in flow chart 1 to measure the density of abnormal cells in the patient’s blood by flow cytometry; When the cells pass through the laser beam, the detector detects the scattered light from the cells or particles; The FSC value is detected by a detector placed in the front; SSC values are detected by multiple detectors placed on the side. By combining forward scattering FSC and transverse scattering SSC data, cells can be grouped according to their size, shape, and complexity. Since the sample cells do not need to be labeled with any fluorescent compound conjugated antibodies, the FSC value and SSC value are two physical characteristics of cell size and particle size obtained by direct sample analysis. Unstained living cells do not suffer any damage and have characteristic light scattering to obtain density values easily [6]. Flow cytometry (FCM) software (Agilent New Cytometer Dx V6B5R3 USA) [7] was used to analyze relative cell size, relative cell particle density, and other techniques to calculate cell density values in the cell bank.

#### 1.2.2 How to obtain pathologically positive blood cells?

The application of “OptiPrepTM “separation fluid (provided by Wuhan Emmetje Technology Co., LTD.) facilitates flotation of individual layer pools from dense load areas through continuous or discontinuous gradients or simple density barriers [8].

Refer to Table 1, according to the different densities of human blood cells, the cell density values in the cell library were calculated by comparative analysis[9], and pathological positive cells with the same density were distinguished.

**Table 1:** Floating density of different blood cells in human body.

| category | Cells | density |
| --- | --- | --- |
| Normal functional cell density | red blood cell | 1.110 |
|  | Eosinophilic | 1.095 |
|  | Neutrophilic | 1.085 |
|  | Lymphoblastic cell | 1.077 |
|  | T lymphocytes | 1.076 |
|  | B lymphocytes | 1.075 |
|  | Lymphocytes | 1.073 |
|  | Natural killer cells | 1.070 |
|  | monocyte | 1.066 |
|  | platelet | 1.030 |
| Pathological positive cell density | MDS | 1.064 |
|  | malignant cell | 1.057 |
|  | SLE | 1.051 |
|  | Plasmodium-infected cell | 1.097 |

1.2.3 The proportion of the separation solution required is determined based on the density measurements of pathologically positive abnormal cells in the blood of different patients. Reference Table 2:

**Table 2:** Reference table for preparation of stratified solutions.

| “OptiPrep™” separation fluid |  |  |
| --- | --- | --- |
| density(g/ml) | solution process%(v/V) | Iodoxanol%(w/V) |
| 1.076 | 35 | 10.4 |
| 1.062 | 25 | 7.4 |
| 1.056 | 20 | 5.9 |
| 1.049 | 15 | 4.5 |

#### 1.2.4 Separation step

According to the experimental design and reference Figure 1, samples from patients with different types of diseases were selected and measured by flow cytometry, and the cell density values of positive cells in the cell bank were calculated and analyzed. We formulated two kinds of layered liquid with high and low density and the density of the target cells in Table 1 according to the operation in Table 2. Using density gradient sedimentation, the only cells of the same type with the density of the separated liquid were densely packed on the interface of the separated liquid, and the upper layer cells were aspirated, and then the density of the separated liquid was adjusted so as to aspirate and expel the upper layer cells with a density smaller than that of the separated liquid.

**Figure 1.**
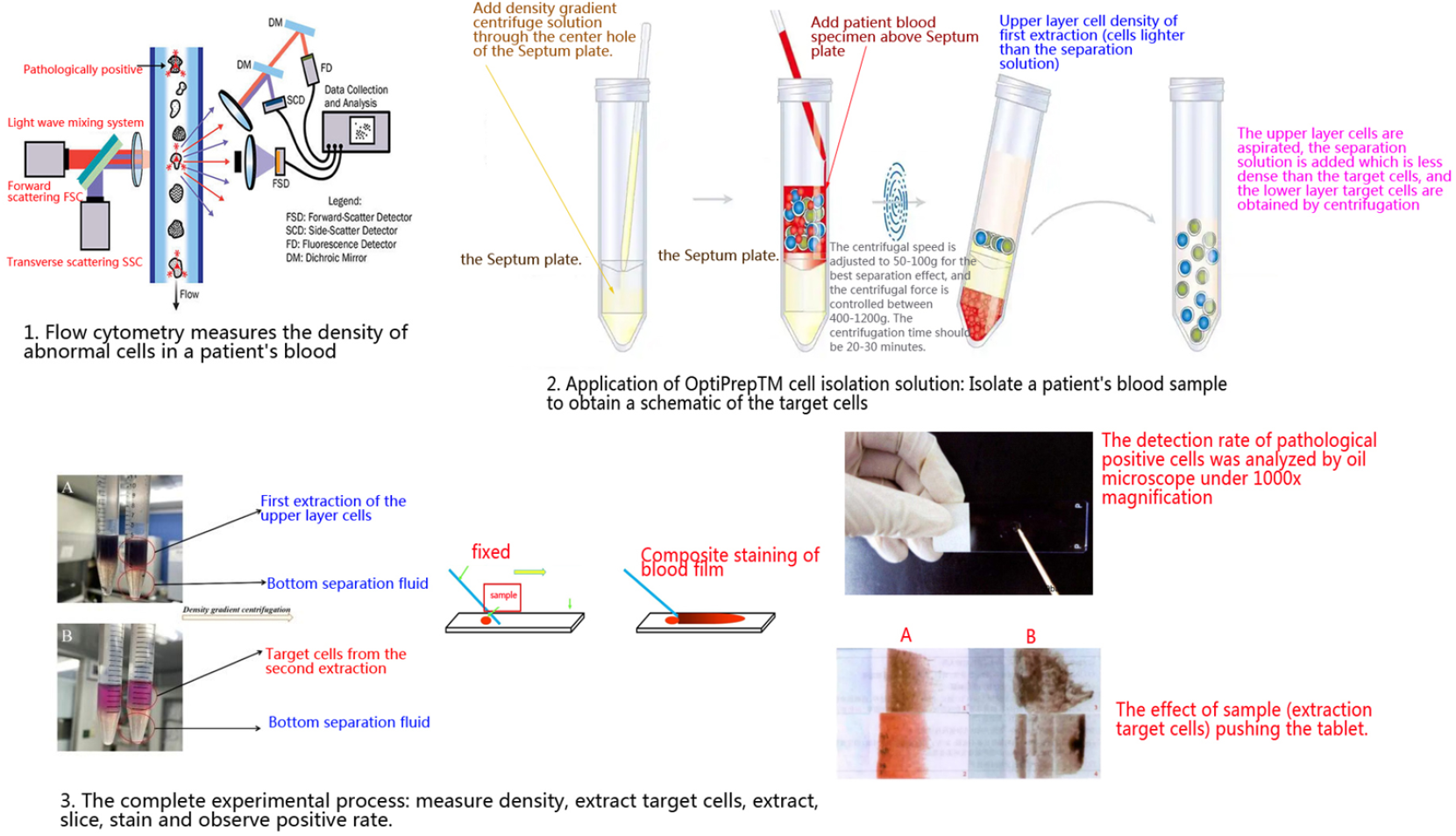
strobe flowchart.

The procedure is shown in Figure 1, where a blood sample is taken and added to a specialized anticoagulant tube. The blood cell density of the patient was calculated by flow cytometry following the steps in the flow chart in Figure 1. The centrifugal speed is adjusted to 50-100g for the best separation effect, and the centrifugal force is controlled between 400-1200g. The centrifugation time should be 20-30 minutes. After adding high density and low density stratified solutions and centrifuging horizontally twice, 98% of the same density cells can be obtained [10]. The pathologically positive abnormal cells with the same separation and purification density were coated on a slide by a special capillary suction method, and the blood film was pushed to air for drying. The cells were first fixed with a drop of methanol solution, then the blood film was stained, and then observed and analyzed with an oil microscope.

#### 1.2.5 Blood membrane cell staining

According to Wright [10], eosin methylene blue is an acidic dye mainly used for microscopic staining, while composite Gilles staining is suitable for staining blood smear specimens, blood cells, plasmodium, Rickettsial, bone marrow cells, immunotumor cells, etc. Please refer to the operation manual for detailed steps.

### 1.3 Comparative Effectiveness of Column Grouping Studies

group I To improve the reliability of the results, we use a manual microscope to observe the diagnostic results. The control group (group II) was also treated with whole blood direct slide staining (automatic slide staining analysis by blood routine line system) to compare the quality and effect of detecting pathologically positive cells.

### 1.4 Statistical comparison methods

Statistical treatment was performed using SPSS version 23, and comparisons between group means were performed using the chi-square test for independent samples; the χ2 test was used for count data. If P<0.05, the difference was statistically significant.

## 2. Results

### 2.1 Diagnostic analysis of observation effect

2.2 Research Group I (Cell Separation, Extraction, and Purification Technology Research Group) and Control Group II (Direct review blood film of same patient)Comparison of positive detection rates of (Table 3):

**Table 3:** Comparison of the total positive rates of the four diseases(%)

\* Chi-square test was used for counting data, and $P < 0.05$ was used for comparison among groups, with statistical significance.
| category | Number of cases | Group 1 | Group 2 | $\chi^2$ | P |
| --- | --- | --- | --- | --- | --- |
| MDS | 33 | 25 (76%) | 9 (27%) | 15.53 | <0.001* |
| malignant cell | 17 | 8 (46%) | 2 (11%) | 6.58 | 0.010* |
| SLE | 29 | 17 (62%) | 7 (33%) | 4.27 | 0.009* |
| Plasmodium-infected cell | 4 | 3 (75%) | 1 (25%) | 4.67 | 0.031* |

## 3. Discussions

Flow cytometry is used to detect and calculate various cell densities in confirmed patient samples. Based on Table 1, the accurate density values of pathological positive cells can be found. OptiPrepTM solution was used in the experiment to prepare two layers of solutions with different densities. The analysis results of the same patient sample in the control group were applied through the blood routine assembly line system to automatically create the rules for microscopic examination and review of blood film thin films, as a comparison of the original data of the control group (Group II).

As shown in Figure 2AB-3AB-4AB-5AB, the target cells of group I were purified, and there were A large number of abnormal positive cells, which improved the accuracy and specificity of diagnosis. Direct smear observation of whole blood samples of group II showed few abnormal positive cells (low detection rate), and the effect was relatively obvious, which was easy to be misdiagnosed and missed diagnosis.

**Figure 2AB-3AB-4AB-5AB:**
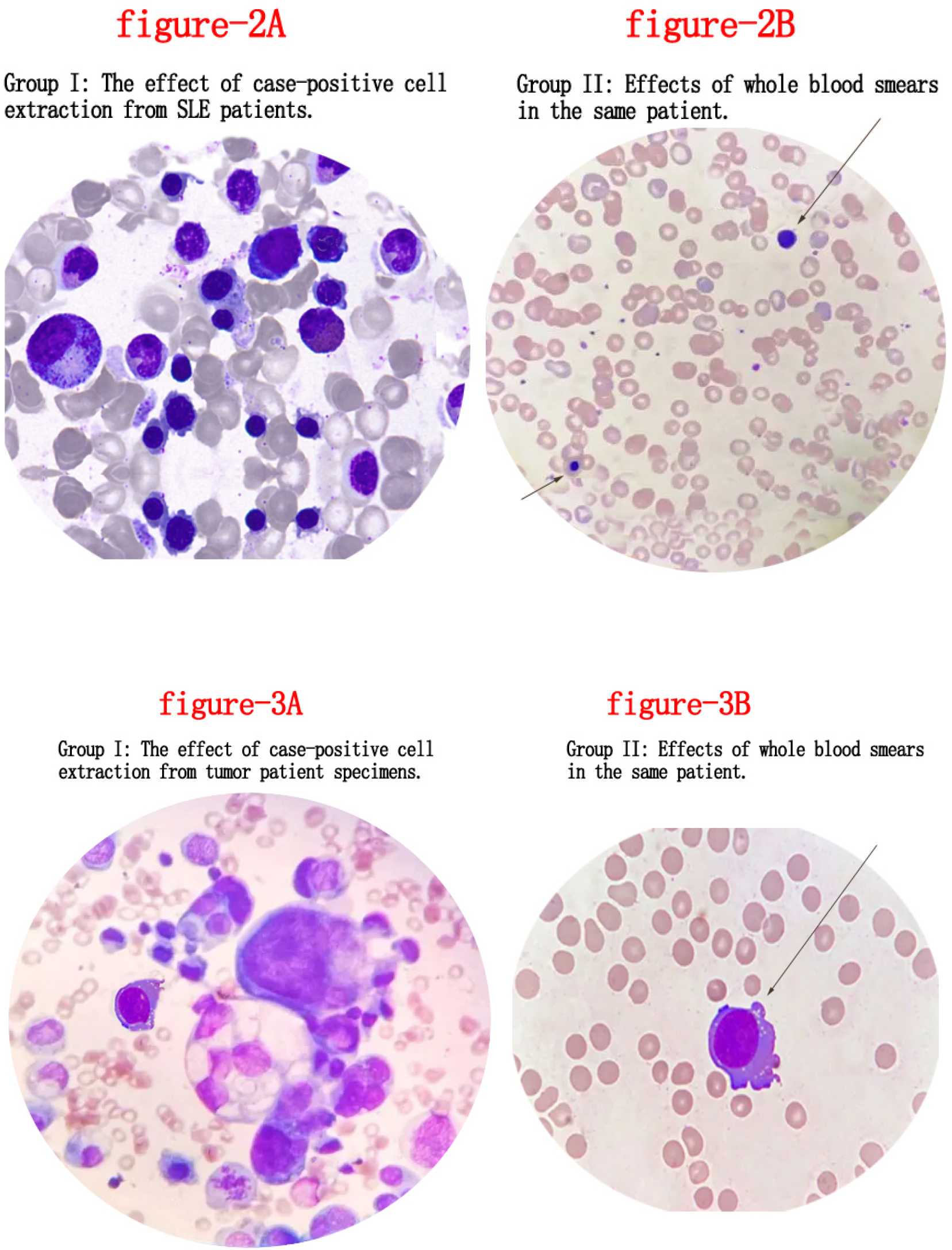
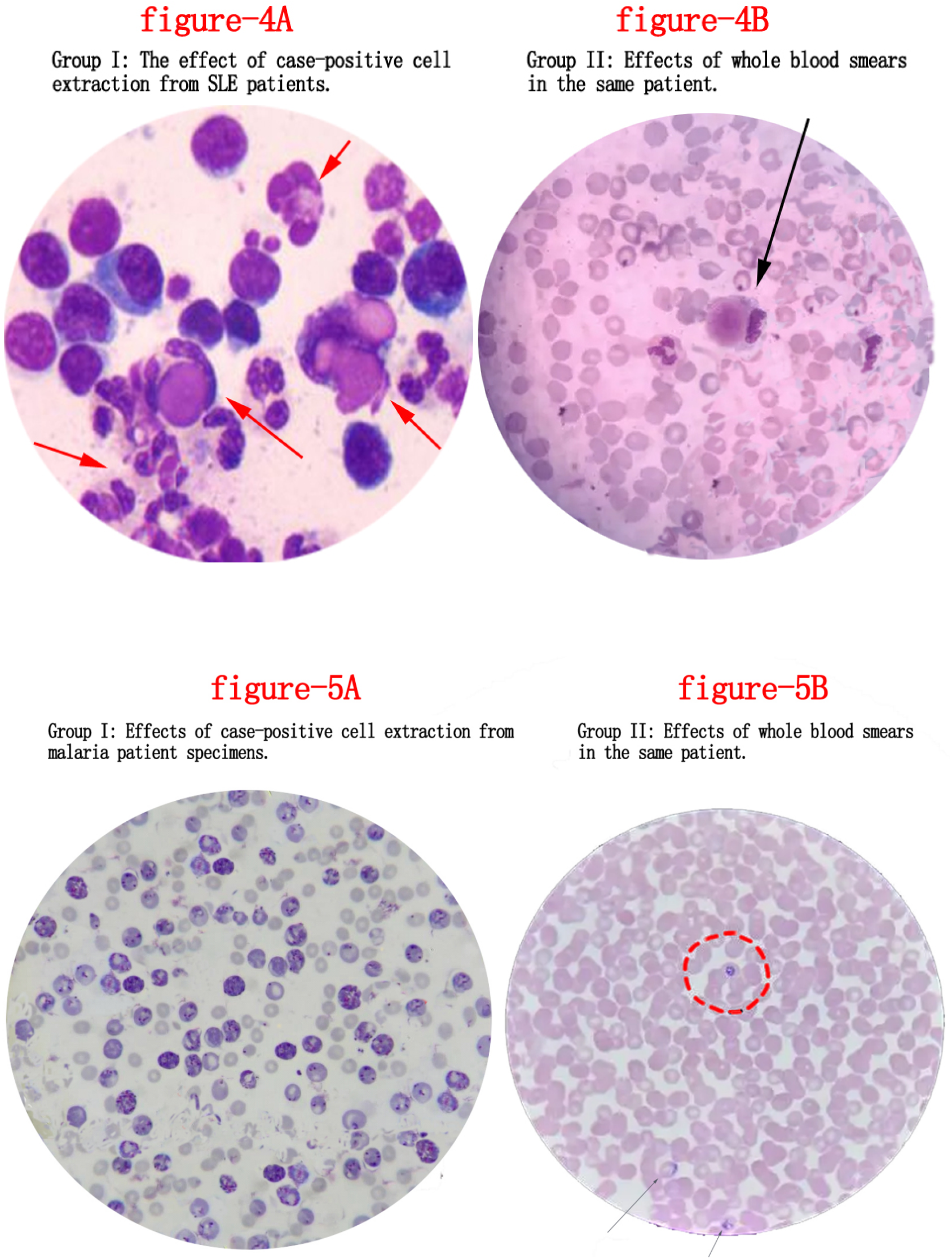
Control effect analysis of positive detection rates of four types of diseases: Study group A (group I) and control Group B (Group II)

Table 3 shows the data before and after comparison of a total of 76 case samples in the two groups. The statistical results of group I showed that the positive test was MDS=76%, malignant tumor cells =53%, lupus cells =62%, and Plasmodium falciparum =75%. In the control group (group II), the positive detection was MDS=27%, malignant tumor cells =11%, lupus cells =33%, and Plasmodium falciparum =25%. Chi-square test was used to compare the mean value of independent samples between groups, and the difference was statistically significant (P<0.05).

Why is the positive detection rate of group I significantly higher than that of group II? This is because more than 98% of pathologically positive cells of the same density were extracted from the target cells for extraction and purification. Comparison (group II) is the automatic push and reading of whole blood samples, which only accounts for a very small part of the patient sample, about 2 microliters, and it is difficult to find the missed diagnosis results in the blood. However, the blood routine assembly line system can automate the efficiency of reprogradation relatively fast and simple, and has advantages[11].

Study group I also failed to achieve a 100 percent detection rate,It may be that the blood samples did not express pathologically positive cells, or there may be errors in experimental data.

In the experimental work, it was found that blood infection diseases first showed abnormal cell lesions, resulting in morphological changes and density reduction [12], which was significantly different from that of normal functioning blood cells. Based on the positive cell density values of different cases, we need to build a rich cell density library to ensure that flow cytometry can analyze and calculate. Characteristic parameters of pathologically positive cells. Therefore, it is easier to find evidence of pathogens, so that the best detection rate can be obtained.

### Advantages and limitations of this study

Important advantages The technology of extracting target cells with separation liquid provides the best detection rate of positive cells, but the limitation is some time-consuming manual operation, and there is no automation to replace the two-step operation of manual cell separation twice.

### Comparison of technical applications with other studies

Compared to the diagnostic techniques in this study, <u>Tehreem Fatim</u>a, indicated that manual microscopy is considered the gold standard. Artificial intelligence needs to be further developed to read and analyze results, which can replace the tedious and time-consuming steps of manually observing results, and there is a growing consensus that it can improve diagnostic efficiency [13]. Corresponding to the morphological characteristics and density index values of tumor cells, Professor <u>Susan Shyu</u>, has studied the effect of selection method for separating human peripheral blood mononuclear lymphocytes, and compared it with our study of flow cytometry for differentiating lymphoma cell density technology to improve the detection rate of similar pathological cells, which can confirm the consistency of the view [14]. <u>Chia-Hsing Liu</u>, studied the cell density of various interstitial fluid, cerebrospinal fluid, body fluid, and body fluid as well as the tissue staining of positive cells extracted from the isolated fluid, and believed that the staining effect had an impact on the correctness of the results of purified and extracted cells [15].

## Conclusions

The density of pathologically positive blood cells can be measured by flow cytometry according to the specimens that need to be reviewed by microscopy as indicated by blood routine, and the target cells extracted can be accurately separated, that is, the same density of pathologically positive blood cells can be collected. Combined with the gold standard method of artificial microscopy, the detection rate was significantly improved, and the occurrence of misdiagnosis and missed diagnosis was reduced. For the difference in cell density of various forms in different types of specimens, it only needs to be distinguished from normal cell density, and abnormal pathological positive cells can be accurately extracted, which can help diagnose the positive detection rate of diseases, which is of great significance and value.

## Data Availability

All relevant data are within the manuscript and its Supporting Information files.

## Notes

### Competing Interest Statement

The authors have declared no competing interest.

### Funding Statement

The author(s) received no specific funding for this work.

### Author Declarations

Sihui People's Hospital, Level 3A Hospital's Medical central laboratory (GuangDong province,CHINA 526200) Medical Ethics Expert Committee of Sihui City Health Management Department Approval Number (ID Number): (Medical Review 2023 No. 61) 2023-061

